# Possible Role of P-selectin Adhesion in Long-COVID: A Case of Recovery After Long-COVID

**DOI:** 10.1101/2023.02.16.23285993

**Authors:** Michael Tarasev, Xiufeng Gao, Marta Ferranti, Aliya U. Zaidi, Patrick Hines

## Abstract

**Background:** Long-term outcomes of severe acute respiratory syndrome coronavirus 2 (SARS-CoV-2) are now recognized as an emerging public health challenge - a condition termed post-acute coronavirus 2019 syndrome (PACS) or Long-COVID. The pathophysiology of Long-COVID remains to be established, several mechanisms in study focus on the role of P-selectin, an inflammation-induced protein expressed by platelets and endothelial cells. Functional P-selectin activity, potentially implicated in COVID-19 and Long-COVID sequelae, was measured for a Long-COVID subject at 68 weeks from the SARS-CoV-2 infection after fully recover from the syndrome. It was compared with the results from the same subject at 20 weeks post-infection, when subject experienced severe Long-COVID symptoms.

**Methods:** Flow adhesion of whole blood or isolated white blood cells to P-selectin (FA-WB-Psel and FA-WBC-Psel) was measured using a standardized microfluidics clinical assay; impedance aggregometry with a collagen agonist was measured using model 590 Chrono-Log impedance aggregometer; standard laboratory assays were performed to evaluate changes in blood chemistries.

**Results:** After recovery from Long-COVID, RBC count and D-dimer remained elevated and other blood chemistry results remained within the normal range as compared to 20 weeks post infection when severe Long-COVID symptoms were present. Total iron and transferrin-iron saturation percentage values that were elevated when symptoms were present, declined to normal range. Whole blood aggregometry results indicate an absence of previously present platelet hyperactivity. FA-WB-PSel that was significantly elevated during Long-COVID (590 ± 260 cells/mm2) was significantly reduced after its symptoms’ resolution (98 ± 38 cells/mm2). However, supplementation of whole blood with crizanlizumab did not result in any measurable inhibition of cell adhesion to P-selectin, similarly with previously reported. Similar to what was observed for the subject when Long-COVID symptoms were present, crizanlizumab, even at a dose 10-fold lower than clinical, induced pronounced inhibition of FA-WBC-Psel when tested in buffer, but not in patient’s own plasma.

**Conclusions:** This report documents the changes in leukocyte adhesion properties for a patient at more than a year from the initial infection after the gradual resolution of Long-COVID symptoms, as compared to when Long-COVID symptoms were present. Recovery from Long-COVID may be associated with normalization of platelet activity, but not necessarily with complete alleviation of endothelial activation. It remains to be determined to what extent changes in leukocyte adhesion to P-selectin may represent a new risk factor for a mechanism driving Long-COVID syndrome.

## Background

A rise in unexplained symptoms following acute coronavirus disease of 2019 (COVID-19) is shown in primary care, where many patients are experiencing prolonged and distressing symptoms which may arise several weeks to months after the exposure to severe acute respiratory syndrome coronavirus 2 (SARS-CoV-2). This condition known as post-acute coronavirus 2019 syndrome (PACS), or post-acute sequelae of SARS-CoV-2 (PAS), is commonly referred to as Long-COVID.

Long-COVID is a complex systemic syndrome associated with substantial morbidity. Despite being included in the ICD-10 classification since September 2020 under the name of post-COVID-19 condition, its definition remains elusive. The preliminary resolution derived through a WHO-led Delphi process, defined Long-COVID as a condition occurring in individuals with suspected or confirmed SARS-CoV-2 infection, typically 3 months after the onset of COVID-19, with atypical chronic symptoms that “last for at least 2 months and cannot be explained by an alternative diagnosis” [1]. Such symptoms may include, among others, extreme fatigue, shortness of breath, headache and migraines, joint and chest pains, brain fogs, loss of smell or taste, hypertension, dizziness on standing, heart palpitations, dermatological complications, hair loss, anxiety and depression [2]. The presence and severity of the symptoms may also fluctuate between individuals and over time. Such heterogeneity of the associated symptoms significantly complicates the assessment of Long-COVID prevalence. PROSPERO study reported that 32 percent of patients experienced extreme fatigue and 22 percent suffered cognitive impairment 2 or more weeks following COVID-19 diagnosis [3]. FAIR Health study reviewed a total of 1,959,982 COVID-19 patients for the prevalence of Long-COVID condition 30 days or more after their initial diagnosis with COVID-19, 23 percent of the patients reported at least one Long-COVID symptom [4]. And a recent meta-analysis review reported that the global prevalence of Long-COVID reaches 49 percent (95% CI, 40-59%) at 120 days after the infection with the two most common symptoms reported being fatigue and memory problems with prevalence of 23 and 14 percent respectively [5].

Persistent and prolonged effects of COVID-19 are often associated with the development of an inflammatory state linked to potentially variegated immunological response [6, 7]. Inflammatory and coagulative sequelae are mediated by selectins including P, E, and L selectin and cell adhesion molecules like ICAM-1, VCAM-1. The current understanding of P-selectin’s role in the pathogenesis of COVID-19 remains to be fully clarified [8]: while some studies did not observe an elevation of P-selectin during the SARS-CoV-2 infection [9, 10], other investigators report significantly elevated P-selectin in intensive care unit patients with COVID-19 disease, compared with healthy controls and COVID-asymptomatic or mildly ill COVID-19 patients [11], with mortality significantly correlated with the elevation of P-selectin [12]. Elevated, predominantly through shedding from activated platelets, levels of soluble P-selectin (sP-sel) had been reported in plasma of COVID-19 patients [9]. P-selectin in platelets interacts with P-selectin glycoprotein ligand-1 (PSGL-1) on leukocytes promoting platelet-leukocyte aggregate formation and release of procoagulant microparticles, and sP-sel levels were suggested as an early marker of thromboembolism [13]. Convalescent COVID-19 patients with no reported Long-COVID symptoms, seems to have levels of VCAM-1, ICAM-1, and P-selectin not significantly different from healthy controls [14]. However, increased levels of platelet-derived (P-selectin+) and leukocyte-derived (CD45+) extracellular particles were reported for COVID-19 patients 30-days post-discharge, indicating the possibility of cellular activation persisting long after the acute phase of the disease.

Pathophysiological mechanisms underlying Long-COVID are a subject of considerable interest but are still far from being fully explained. Evidence on why persistent symptoms occur is still limited and available studies are heterogeneous. Apart from long-term organ damage, including possibly damage to the autonomic nervous system, many hints suggest that diverse Long-COVID symptoms could arise from different specific mechanisms following acute illness [15]. Possible mechanisms that could contribute to the variability of the symptomatology include immune dysregulation, auto-immunity, endothelial dysfunction, occult viral reservoirs, as well as coagulation activation. As opposed to the direct effects of the virus itself, the appearance of lasting Long-COVID symptoms is believed to be due to prolonged or delayed immune response, possibly through facilitation of an ongoing inflammatory processes and an organ-specific post-SARS-CoV-2 infection sequelae [15-17].

We previously reported platelet hyperactivity associated with increased P-selectin activity and seemingly with alteration in functionality of PSGL-1 leukocyte receptor for a Long-COVID patient, as compared to post-COVID asymptomatic subject. Such alteration of cellular reactivity was associated with severe and prolonged, up to 5 months after COVID-19 resolution, symptomatology, which included many of the typical symptoms associated with the Long-COVID condition. In the present work, we report the changes observed after the patient’ recovery (at 68 weeks) from Long-COVID and complete amelioration of associated symptoms.

## Methods

### Study Subjects and Sample Collection

Convalescent COVID-19 subject, with previous Long-COVID symptoms (LC) was recruited according to the protocols FF-RBC-001 and FF-RBC-003v2 approved by Institutional Review Board of the Institute for Regenerative and Cellular Medicine. The blood draws were obtained via venipuncture after a 12-h fast. Whole blood was collected into K2-EDTA tube for CBC data, 3.2% sodium citrate tube for aggregometry, flow adhesion assays, and D-dimer, serum separation tubes for interleukins, COVID-19 antibodies, calcium, iron, TIBC and ferritin panel. Flow adhesion and aggregometry assays were performed in-house, with other assays performed by Quest Diagnostic Lab. Reference ranges are those provided by the laboratory unless specified otherwise. The subject consented to take part in the present study and for the results to be published.

### Flow Adhesion Assays

Flow adhesion of whole blood to P-selectin (FA-WB-Psel) and Flow adhesion of white blood cells to P-selectin (FA-WBC-Psel) were conducted as described previously [18]. Briefly, isolated white blood cell (i-WBC) suspensions were prepared according to a standardized protocol (HetaSep™, StemCell Technologies). Whole blood samples (1:1 diluted with HBSS buffer) or i-WBC (5× 10^6^ cells/mL) were perfused through P-selectin-coated microfluidic channels at 1 dynes/cm^2^ for 10 or 6 minutes respectively, washed to eliminate non-adhering cells, with resultant adhesion quantified manually by an independent trained observer to generate an adhesion index (cells/mm²) using a previously described protocol [19]. For drug treatment conditions, samples were incubated for 5 minutes with crizanlizumab at the final concentrations as required before assessment by the flow adhesion assay. Crizanlizumab was from Creative Biolabs, Shirley, NY; P-selectin was a disulfide-linked homodimer from R&R systems, Minneapolis, MN.

### Impedance Aggregometry

Platelet aggregation in whole blood samples was tested with an impedance aggregometer (Model 590; Chrono-Log Corporation, Havertown, PA) and analyzed using Aggro/Link®. Whole blood sample was transferred at room temperature and tested within 4 hours of the blood draw. The sample was split into control (1% DMSO treated) and aspirin treatment (100µM). Briefly, after a stable baseline (steady state) had been established, the agonist collagen (Chrono-Par, Chrono-Log Corporation, Havertown, PA) was added to the sample to a final concentration of 2µg/mL, and the aggregation was monitored for 10 minutes. Instrument normal control reference values (Table 3) are for normal donor with no previous exposure to COVID-19 tested same as post-COVID subjects.

### Statistical Analysis

The data is presented as mean ± standard deviation (mean ± SD) with Student t-test paired or non-paired as appropriate used for assessment of statistical significance of the differences. Results were deemed significant for comparisons where two-tailed *p* < .05.

## Results

### Clinical Sequelae

LC is a 26–30-year-old female with no history of chronic medical conditions, was not on any prescription medication, and had no prior health complaints. She was diagnosed with SARS-CoV-2 by PCR and presented with COVID-19 symptoms one week following a known exposure, including coughing, fever, difficulty breathing, progressively worsening fatigue, brain fog, loss of taste and smell, and headache. LC was not hospitalized, managing symptoms at home. At most severe (10-12 days after the infection), she was on bed rest, not responsive, refusing food or drink. The symptoms started to alleviate when the fever was broken and gradually receded over the course of another week. After the resolution of the infection, LC experienced a wide range of symptoms including, extreme fatigue, brain fog, hair loss, headaches, and migraines, which are typical for those reported in Long-COVID cases. While some symptoms started to alleviate, many of them remained severe beyond 5 months after the infection [20]. At 68 weeks post-infection, she considered herself recovered and does not report any lingering symptoms. Her energy level was up to about pre-SARS-CoV-2 infection levels, and she no longer suffered from migraines and brain fog.

### Clinical Chemistry

During Long-COVID, clinical chemistry results were predominantly within the normal ranges. The exceptions were: elevated D-Dimer level, slightly elevated RBC count combined with the decreased total iron and transferrin-iron saturation percentage values on the background of normal total iron binding Capacity (TIBC). After recovery from Long-COVID, total iron and transferrin-iron saturation percentage values were within the normal range, however the slight increase in RBC count and D-Dimer value remained unchanged as compared to 20 weeks post infection, when severe Long-COVID symptoms were present (Table 1).

**Table 1.**

|  | Long-COVID subject (female) |  |  | Reference range<sup>1</sup> | Average reported values (mean ± SD) |
| --- | --- | --- | --- | --- | --- |
| Weeks past the resolution of SARS-CoV-2 infection | 11 weeks | 20 weeks | 68 weeks |  | During active COVID-19 |
|  | Long-COVID Symptoms |  | Recovered |  |  |
| Complete blood count (CBC) |  |  |  |  |  |
| WBC, bil/L | 7.8 | 9.9 | 8.0 | 3.3 – 10.7 | 6.0 ± 2.0 |
| RBC, tril/L | 4.92 | ⬆ 5.26 | ⬆ 5.25 | 3.8 - 5.1 | 4.2 ± 0.4 |
| Hemoglobin, g/dL | 13.4 | 14.2 | 14.2 | 12.1 - 15.0 | 13 ± 1.1 |
| Hematocrit, % | 41.5 | 43.6 | 44.3 | 35.4 - 44.2 | 41.6 ± 2.7 |
| MCV, fL | 84 | 82.9 | 84.4 | 80 - 100 | 91.5 ± 5.2 |
| MCH, pg | 27 | 27 | 27 | 27 - 33 | 30.6 ± 2.1 |
| MCHC, g/dL | 32 | 32.6 | 32.1 | 32.0 - 35.0 | 33.3 ± 1.1 |
| RDW-CV, % | 14 | 13 | 14 | 11 - 15 | 14.3 ± 1.7 |
| Platelets, bil/L | 318 | 310 | 346 | 150 - 400 | 183 ± 39 |
| MPV, fL | N/A | 10.2 | 10.5 | 7.5 - 12.5 | 7.8 ± 4.2 |
| Metabolic Panel and electrolytes |  |  |  |  |  |
| Ferritin, ng/mL | N/A | 26 | 24 | 11-172 | ⬆⬆ 1018 ± 533 |
| Iron, Total, mcg/dL | N/A | ⬇ 35 | 78 | 40 - 190 | 150 ± 55 |
| Iron Binding Capacity (TIBC), mcg/dL | N/A | 358 | 326 | 250 - 450 | 390 ± 53 |
| Transferrin-iron saturation percentage (TSAT), % | N/A | ⬇ 10 | 24 | 16 - 45 | 29 ± 10 |
| Other parameters |  |  |  |  |  |
| D-Dimer, mcg/mL FEU | N/A | ⬆⬆ 1.02 | ⬆⬆ 1.01 | <0.50 | ⬆⬆ 1.2 ± 1.4 |

Values above the normal reference range are indicated by ↑ for moderate increase and by ↑↑ for large increase, and values below the normal reference range indicated by ↓ for moderate increase and by ↓↓ for large increase. All other values are within the reference range. Abbreviations: WBC, white blood cell; RBC, red blood cell; MCV, mean corpuscular volume; MCH, mean corpuscular hemoglobin; MCHC, mean corpuscular hemoglobin concentration; RDW-CV, red cell distribution width; MPV, mean platelets volume.
<sup>1</sup>The reference ranges are those provided by Quest Diagnostics.

### Flow Adhesion on P-selectin

#### Whole Blood

Flow adhesion of whole blood on P-selectin (FA-WB-PSel) that was significantly elevated during Long-COVID (590 ± 260 cells/mm^2^) was significantly reduced after symptoms’ resolution (98 ± 38 cells/mm^2^). However, FA-WB-PSel value remained elevated as compared to the values anticipated in normal subjects when no significant inflammation processes were present. The subject did not report any symptoms or trauma events that could have resulted in the elevation of FA-WB-PSel levels due to other unrelated events. Supplementation of whole blood with crizanlizumab did not result in any measurable inhibition of cell adhesion to P-selectin, similarly to what was observed previously during the Long-COVID syndrome. Moreover, elevated adhesion was observed at low doses of the drug (Figure 1).

**Figure 1:**
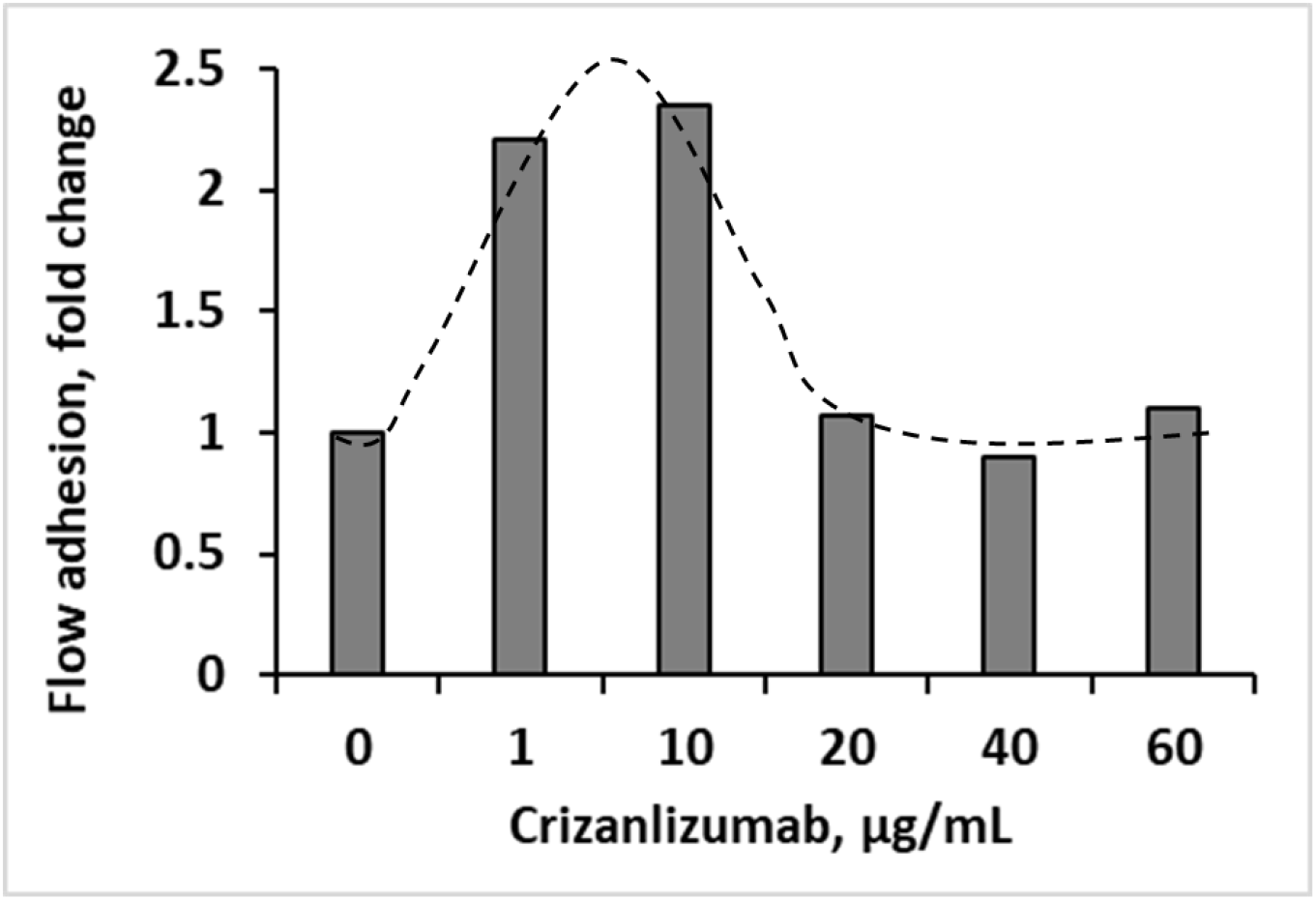
Flow adhesion on P-selectin substrate with whole blood sample diluted 1:1 with HBSS buffer supplemented with crizanlizumab. Trend line is for an illustrative purpose only. Adhesion is shown as a fold change relative to the no-drug baseline.

#### Isolated White Blood Cells

After the symptom’s resolution, flow adhesion of isolated white blood cells (i-WBC) to P-selectin (FA-WBC-Psel) in HBSS buffer was significantly reduced, to 628 ± 12 cells/mm^2^, from the observed 1490 ± 30 cells/mm^2^ when Long-COVID symptoms were present. Supplementation of i-WBC with crizanlizumab showed nearly complete (down to 99% at high dose) inhibition of cell adhesion. Similar to what was observed for the subject when Long-COVID symptoms were present [20], such strong adhesion inhibition occurred even at 1 µg/mL of crizanlizumab, which is ten times lower than the standard clinical drug dose (Figure 2).

**Figure 2.**
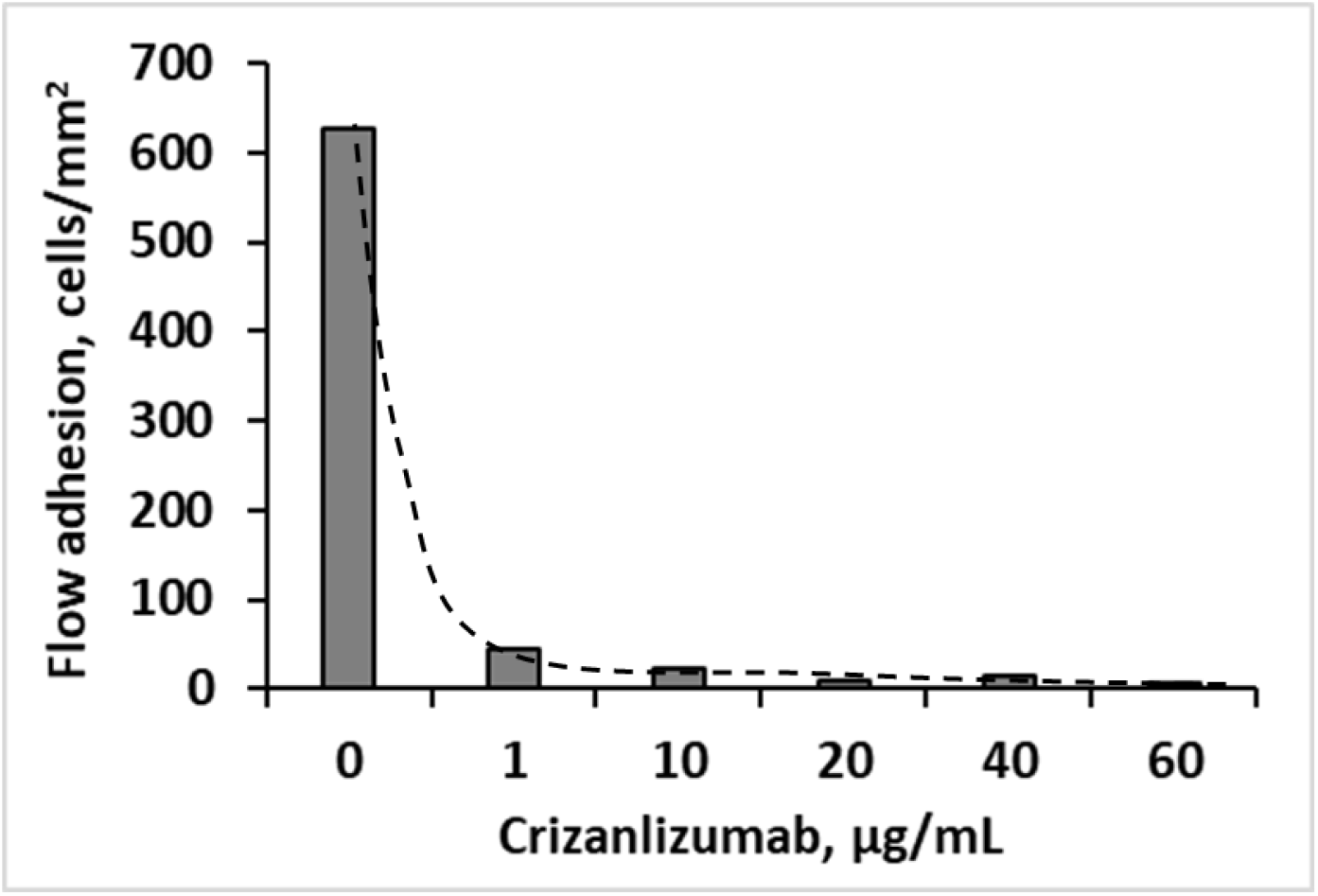
Isolated white blood cell flow adhesion on P-selectin substrate HBSS buffer supplemented with crizanlizumab. Trend line is for an illustrative purpose only.

When suspended in autologous platelet poor plasma (PPP), i-WBC were less adhesive than in HBSS buffer (flow adhesion of 245 ± 30 cells/mm^2^ as compared to 628 ± 12 cells/mm^2^ in HBSS). This 2.6-fold reduction in adhesion is smaller, if comparable to the 3.8-fold reduction in adhesion observed when Long-COVID symptoms were present. Incubation with crizanlizumb resulted in reduction in adhesion in a drug dose-dependent manner. About 40 percent inhibition of adhesion was achieved at 10 µg/ml, that corresponds to the drug clinical dose, with the maximum inhibition of about 75 percent observed for the highest used crizanlizumb dose of 60 µg/ml (Figure 3). During Long-COVID, incubation of subject’s i-WBC in PPP with 10 µg/ml crizanlizumb resulted in a more pronounced inhibition of adhesion (about 65 percent); changes in adhesion at higher drug doses were not tested.

**Figure 3.**
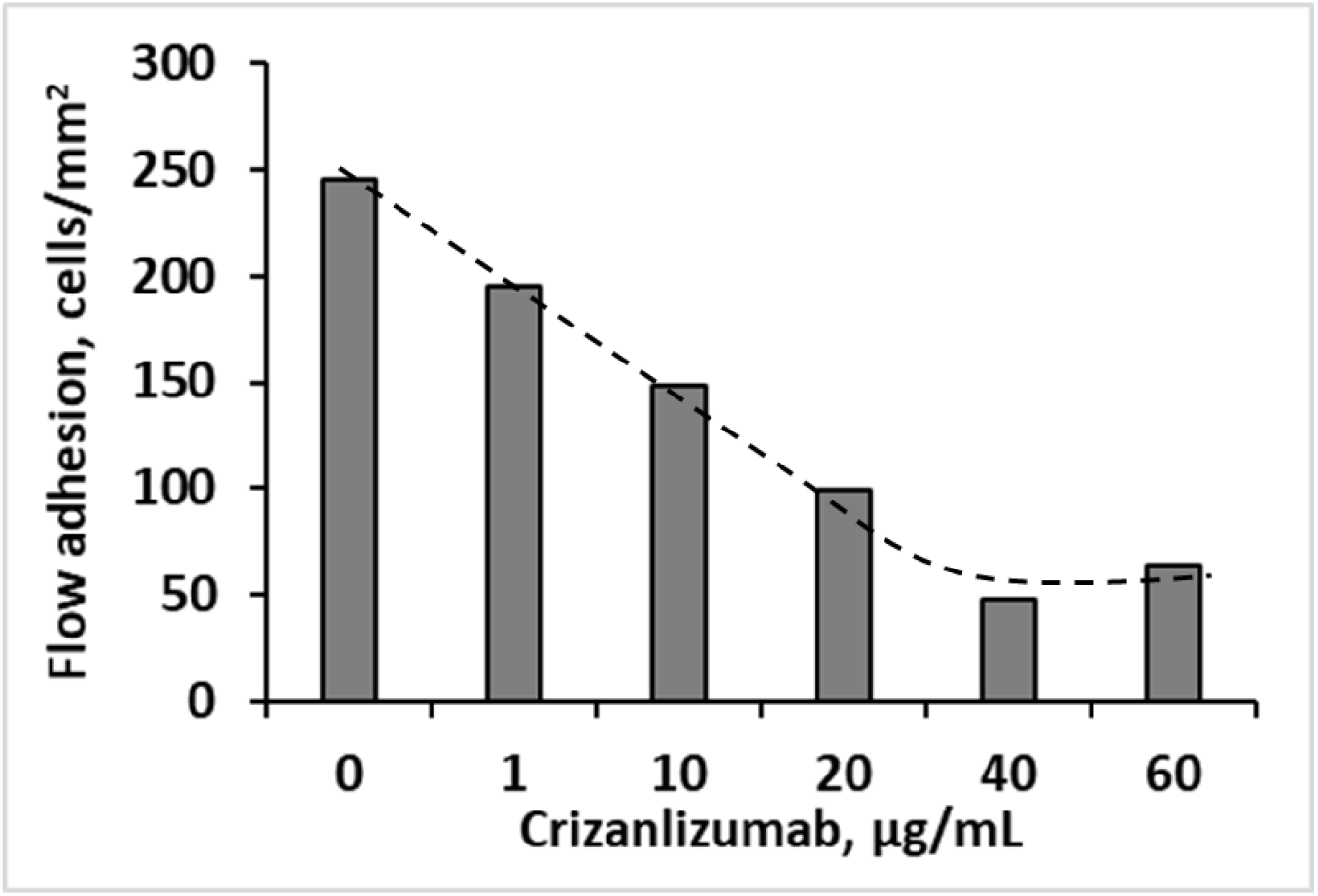
Flow adhesion on P-selectin substrate of isolated WBC subject’s own platelet poor plasma supplemented with crizanlizumab. Trend line is for an illustrative purpose only.

### Whole Blood Aggregometry

Whole blood impedance aggregometry showed that maximum amplitude, area under the curve (AUC) and maximum rate that were elevated during Long-COVID when symptoms were present, decreased to normal control values after the symptoms’ resolution (Table 2). Lag time remains unaffected regardless of the presence or absence of Long-COVID symptoms. Inhibition of activity induced by antiplatelet acetylsalicylic acid (ASA) that was found to be more pronounced in Long-COVID subject as compared to the normal control, was smaller and closer to normal control values.

**Table 2.** Whole blood impedance aggregometry

|  | No ASA |  |  |  | + ASA<sup>1</sup> |  |  |  |
| --- | --- | --- | --- | --- | --- | --- | --- | --- |
|  | Internal<br>healthy<br>control | Long-Covid subject<sup>2</sup> |  | Significance<sup>3</sup> | Internal<br>healthy<br>control | Long-COVID subject<sup>2</sup> |  | Significance<sup>3</sup> |
|  |  | 20 weeks | 68 weeks |  |  | 20 weeks | 68 weeks |  |
| Amplitude,<br>Ohm | 4.5 | 11.4 ±<br>0.2 | 5.4 ± 0.2 | $p < 0.05$ | 2.3 | 1.5 ± 0.2 | 3.7 ± 0.2 | $p < 0.05$ |
| Rate,<br>Ohm/min | 1.5 | 8.1 ± 0.2 | 0.9 ± 0.2 | $p < 0.05$ | 0.5 | 0.45 ±<br>0.05 | 0.7 ± 0.1 | ns |
| Lag time,<br>min | 1.5 | 1.4 ± 0.1 | 1.5 ± 0.2 | ns | 3.5 | 2.3 ± 0.1 | 2.2 ± 0.2 | ns |
| AUC (at 10<br>min) | 35 | 80 ± 7 | 31 ± 6 | $p < 0.05$ | 15 | 9 ± 3 | 22 ± 6 | $p < 0.05$ |

<sup>1</sup>Supplemented with 100 µM aspirin<sup>2</sup>As reported previously [20]<sup>3</sup>Significance: t-test comparison between the results obtained at 20 weeks (5 months) when severe Long-COVID symptoms had been present and 68 weeks (17 months) when symptoms had been resolved.
Note: Experimental error was evaluated based on signal to noise ratios of experimental data.

## Discussion

Recovery from Long-COVID is a long process that can take many months. Abnormal fatigue, lack of muscle strength, and impaired cognitive functioning, among other symptoms, was reported for Long-COVID patients 10 months after recovery from the infection [21]. A recently published study tracked antinuclear/extractable-nuclear antibodies in patients with Long-COVID and reports a correlation between antibody values and prevalence of inflammation and symptoms like fatigue and respiratory problems as far as 12 months after COVID-19 [22]. A 2-year retrospective data analysis of health records of approximately 89 million patients indicated that while increased risks of mood and anxiety disorders was transient, the risks of neurological complications like psychotic disorder, cognitive deficit, dementia, and epilepsy remained elevated, with the hazard ratio for some increasing over time up to 2 years of study observation [23].

We previously reported a case of Long-COVID subject with severe multifaceted symptom presentation up to 5 months after the resolution of the initial SARS-CoV-2 infection. This subject presented platelet hyperactivity that was associated with increased P-selectin activity and seemingly with alteration in the functionality of PSGL-1 leukocyte receptors. These features were not observed in an asymptomatic post-COVID subject [20]. At 20 weeks, when severe Long-COVID symptoms were present, crizanlizumab, an anti-P-selectin monoclonal antibody, was found to be less potent in whole blood based on cell adhesion to immobilized P-selectin.

At 68 weeks, with apparent symptoms of Long-COVID reported by the subject as being resolved, whole blood aggregometry results indicate an absence of previously present platelet hyperactivity with substantially reduced WB-FA-PSel. It was suggested previously that platelet activation in COVID-19 may be due to platelet interaction with the infected endothelium and/or because of the cytokine storm associated with SARS-CoV-2 infection [24]. If that is indeed the case, recovery from Long-COVID may be associated with normalization of platelet activity, but not necessarily with complete alleviation of endothelial activation.

Interestingly, WB-FA-PSel was elevated when the blood sample was incubated with lower doses of crizanlizumab, without a measurable inhibition of P-selectin adhesion even at very high drug dose. We previously suggested that the failure of P-selectin inhibitor to reduce cell adhesion to immobilized substrate may be due to its interaction with the native P-selectin present in the blood sample (e.g., soluble P-selectin in plasma or expressed on activated platelets). If significant, binding of P-selectin antagonist to blood P-selectin could reduce the amount of antagonist available for blocking immobilized P-selectin in the assay and by extension, membrane-bound P-selectin on endothelial cells.

As only the lectin and epidermal growth factor domains are necessary for P-selectin interaction with PSGL-1 [25], monomeric sP-sel is capable of binding to high affinity ligands on leukocytes. Despite soluble protein lower avidity for leukocytes as compared with its oligomeric transmembrane form [26], the implication seems to be that by blocking at least some binding sites, sP-sel can be reducing apparent leukocyte adhesion to membrane bound, oligomeric P-selectin. Thus, competitive binding of sP-sel with P-selectin antagonist like crizanlizumab, would decrease the number of blocked P-selectin counter-receptors on leukocytes, and if the drug concentration is sufficiently low, lead to an apparent increase of cell adhesion on immobilized (or membrane-expressed) P-selectin. Alternatively, P-selectin antagonist could be disrupting the interactions between platelet-expressed P-selectin and leukocyte PSGL-1, and at sufficiently low concentrations, similarly resulting in elevation of cell adhesion due to more cells and/or higher levels of PSGL available for down-stream interaction with immobilized protein. Additional work is required to validate and potentially differentiate between these scenarios of P-selectin antagonist behavior on the background of platelet activation and high sP-sel.

## Conclusions

This report documents the changes in leukocyte adhesion properties for a patient at more than a year from the initial infection after the gradual resolution of Long-COVID symptoms, as compared to the same patient during Long-COVID. Despite significant reduction in platelet hyperactivity observed when the symptoms of Long-COVID are resolved, changes in i-WBC adhesion to P-selectin in the presence of P-selectin antagonist remain effectively unchanged from when severe Long-COVID symptoms were present. Recovery from Long-COVID may be associated with normalization of platelet activity, but not necessarily with complete alleviation of endothelial activation. It remains to be determined to what extent changes in leukocyte adhesion to P-selectin may represent a new risk factor for or a mechanism driving Long-COVID syndrome.

## Data Availability

All data produced in the present study are available upon reasonable request to the authors

## Conflict of interest statement

M. Ferranti and A. Zaidi are employees, and X. Gao, P. Hines and M. Tarasev are employees and shareholders of Functional Fluidics Incorporated, a company developing and commercializing assays for assessment of blood cell functions.

## Notes

### Funding Statement

This study was internally funded by Functional Fluidics Inc, a company that develops assays for assessment of functional properties of blood cells.

### Author Declarations

Ethical approval for the study was under the protocols FF-RBC-001 and FF-RBC-003v2 approved by Institutional Review Board of the Institute for Regenerative and Cellular Medicine.

